# Laboratory study to investigate the sensitivity and specificity of a diagnostic dressing to detect burn wound infection

**DOI:** 10.1101/2021.07.21.21260922

**Authors:** N.T. Thet, A.T.A. Jenkins, J.D. Mercer-Chalmers, K. Coy, S. Booth, D. Collins, S. Bremner, F. Spickett-Jones, R. Greenwood, A.E. Young

## Abstract

Burn Wound Infection (BWI) is difficult to define and detect before it manifests with clear clinical symptoms. In this paper, an *ex vivo* study of a prototype BWI detecting wound dressing is reported. Consenting patients with burns were recruited from four burns services in the United Kingdom, their burn infection state recorded at time of recruitment and retrospectively following treatment. Their wound dressings were used as a source of inoculating bacteria to create an *ex vivo* biofilm model in the laboratory with reasonable fidelity to the original microbial state of their wound. The prototype infection detecting wound dressing, which responds to cytolytic toxins secreted by bacteria, was placed on the *ex vivo* biofilm and the response of the dressing correlated with the clinical decision on the patient’s wound infection state. The study illustrated a number of broader issues with clinical BWI diagnosis, notably the absence of objective diagnostic criteria: a ‘reference standard’ for BWI. The absence of such a reference standard made analysis of the relationship between the dressing response and BWI diagnosis challenging, however a point estimate of 68% sensitivity from the study suggests the potential future utility of using a sensor which detects secreted bacterial virulence factors to assist in BWI diagnosis.

## Introduction

Burn wound infection (BWI) is a complex multi-factorial state which is difficult to define and diagnose. It occurs as a complication in 10-20% of patients with small area burns and in 75% of patients with larger area burns.^1, 2^ BWI results in delayed wound healing, increased permanent scarring and in systemic sepsis if untreated, with associated risks of mortality. The standard method of diagnosing infection, based on swabbing wounds for culture and identification of resultant bacterial colonies, is time-consuming and will not inform clinicians of the bacterial potential to cause harm to the wound or patient. This lack of an accurate point of care (PoC) test forces clinicians to diagnose BWI by means of a combination of judgement and experience, using patient symptoms, signs of inflammation and non-specific laboratory tests. The consequence of this diagnostic gap is that patients with a suspected BWI undergo potentially unnecessary dressing changes, negatively impacting wound healing, and are often pre-emptively treated with systemic antibiotics, without a definitive diagnosis. The ability to provide a more accurate diagnosis at PoC, would result in better outcomes for patients, a reduction in the need for unnecessary antibiotic prescriptions, along with associated reduction in bacterial resistance to antibiotics and in healthcare costs.

To develop a PoC diagnostic for BWI requires an understanding of when a potentially infected wound requires treatment; the state *immediately prior* to clinical infection. This requires knowledge of the wound infection pathway from injury, through to contamination, colonisation and eventually clinically-relevant wound infection. Critical to this, is an appreciation that whilst wound infection requires the presence of pathogenic bacteria, the reverse is not necessarily true: *the presence of bacteria in a wound does not necessarily mean it is infected*. This fact creates challenges when designing a sensor for detecting a clinical infection that requires treatment, with many of the currently used classical approaches to clinical microbiology - which look for microbes (typically bacteria or yeast) – being unlikely to exhibit prognostic efficacy. During the process of wound infection, bacteria attach and colonise a wound within seconds of the skin being breached by the burn. Bacteria in the air and surrounding skin will enter the wound and grow in numbers. The point at which the density of bacteria becomes sufficiently high such that Quorum Sensing (QS) pathways become activated, is considered the point at which clinical wound infection is established.^3^ QS is the molecular genetic mechanism by which bacteria detect their local cellular density, allowing them to switch on cytotoxic virulence factor secretion.

Two of the authors of this paper have developed a prototype ‘smart dressing’ technology to diagnose BWI without dressing removal and at point of patient presentation to medical care. The dressing responds to virulence factor secretion from predominant burn wound pathogens (*Staphylococcus aureus, Pseudomonas aeruginosa, Enterococcus faecalis*) by releasing an indicating dye suspended in nano-vesicles within the dressing. Colour change is triggered at a bacterial concentrations two orders of magnitude lower than their cytotoxicity threshold (level at which treatment is necessary) providing early warning before tissue damage can occur, whilst distinguishing invasive infection from low-level background colonisation. The colour change alerts health professionals, by indicating true wound infection at PoC. The aim of this diagnostic accuracy study is to evaluate the prototype PoC device for sensitivity and specificity to clinically relevant BWI. The study was undertaken using patient samples from infected and non-infected wounds. A key challenge with assessing the diagnostic utility of an infection diagnostic is the lack of a reference standard.^4^ As there are no agreed and objective indicators for diagnosis of BWI at PoC, validating a technology has to be based on clinician judgement and retrospective diagnosis based on patient signs.

## Methods

This study followed a pre-agreed protocol and was registered on the ISRCTN, registration number: 13825483. It was approved by the South West - Cornwall & Plymouth Research Ethics Committee: reference 16/SW/0238.

### Trial design

The *EVIDEnT* (Ex Vivo Infection Detection) study was a four-site pragmatic, single-blind, prospective, observational laboratory study. The aim was to assess the sensitivity and specificity of a prototype *smart dressing* technology for detection of clinically-relevant burn wound infection (BWI). BWI was determined by clinical consensus based on a reference standard comprising three signs of wound infection. Laboratory staff assessing dressing ‘switch-on’ were blinded to patient condition.

### Study objectives

- To define the reference standard for diagnosis of BWI.
- To compare infection detection smart dressing technology with a clinical reference standard to estimate sensitivity and specificity for the detection of BWI.
- To determine true positive rate linking technology ‘switch-on’ to clinical infection as determined by the reference standard.
- To determine true negative rate linking technology ‘remains switched-off’ in response to no clinical infection.
- To determine false positive and negative rates, sensitivity and specificity with 95% confidence intervals for all percentages.
- To determine test accuracy calculated using the above data.

### Population

Participants included those able to consent (with parental consent if < 16 years of age) with burns of any size, any severity and any cause. Participants were recruited from 4th October 2016 – 30th April 2018.

#### Wound Infection Group

Children (of more than 4 weeks of age) and adults (of less than 100 years of age) with diagnosed BWI, not on antibiotics for more than 24 hours, presenting to the University Hospitals Bristol NHS Foundation Trust, Queen Victoria Hospital NHS Foundation Trust, East Grinstead, Chelsea and Westminster Hospital NHS Foundation Trust or North Bristol NHS Trust, Bristol.

#### Control Group

Children and adults presenting to the above sites at more than 48 hrs after burn injury, but before healing and without a diagnosed BWI. The control participants must not have been prescribed antibiotics. Participants remained in the control group providing that they had no diagnosed wound infection by a 14-21-day follow-up.

If a control patient came back to hospital with a diagnosed infection, the patient’s first set of swab results were excluded from the calculation of sensitivity and specificity. Samples collected after infection was diagnosed were included in the wound infection group.

##### Reference standard

Clinically relevant BWI was determined retrospectively by senior clinicians using an agreed reference standard comprising three burn wound symptoms and signs: increased local pain, increased wound temperature and peri-wound redness. The use of these signs was supported by earlier consensus work on indictors for the diagnosis of burn wound infection.^5^

### Outcome Measures

- Technology ‘switched-on’ in response to diagnosed BWI.
- Technology remained ‘switched-off’ in response to no BWI.

#### Follow-up

All participants were followed-up in clinic or by telephone 14-21 days after presentation for a retrospective diagnosis of wound infection or clarification of control status. Those patients who were lost to follow-up were excluded from the study. Patients whose wound samples did not arrive in the laboratory in perfect condition were also excluded.

#### Wound sample management

patient burn wound exudate, biofilm, wound debris and the discarded wound dressing were collected and sealed inside sterile bags or universal containers. One swab was sent for routine analysis (microscopy, culture and sensitivity) in standard swab transfer medium to each of the participating hospital sites’ microbiology departments. All other routine clinical data were recorded as per standard care.

#### Assessment of ‘switch-on’

The University of Bath laboratory staff were blind to patient condition (presence or not of BWI). The samples were assessed against the infection detection technology in terms of ‘switch-on’ (colour change), ‘no switch-on’ or inconclusive. Results were determined by two separate scientific personnel independently and confirmed by photograph. If the two scientific personnel rated the dressings differently, a third person rated the dressing for switch on (or not).

#### Prototype dressing technology

The design and construction of the prototype smart wound dressing technology is detailed in a previous publication.^6^ The dressing consists of a 4 mm thick, 3 cm x 3 cm cryo-crosslinked polyvinyl-alcohol gel embossed with 4 mm diameter wells 0.5 mm deep on the wound side only (figure 1). The wells are filled with a soft agarose gel containing phospholipid vesicles (liposomes) containing a combination of lipids and fatty acids. Inside the vesicle is a high concentration (50 mM) aqueous solution of carboxyfluorescein. At this concentration, the fluorescence of carboxyfluorescein is self-quenched and the dye appears a dull orange colour. The gel ensemble is itself covered in a silicone gel film (Meopore®) which would be used to adhere the dressing to the skin. The vesicles themselves are sensitive to secreted cytolytic bacterial virulence (present in clinically-relevant BWI), which are ruptured, releasing their carboxyfluorescein payload into the surrounding hydrogel. The resultant dilution of the dye causes an intense ‘switch on’ of fluorescence, easily observed by eye as a bright green colour (figure 1).

**Figure 1:**
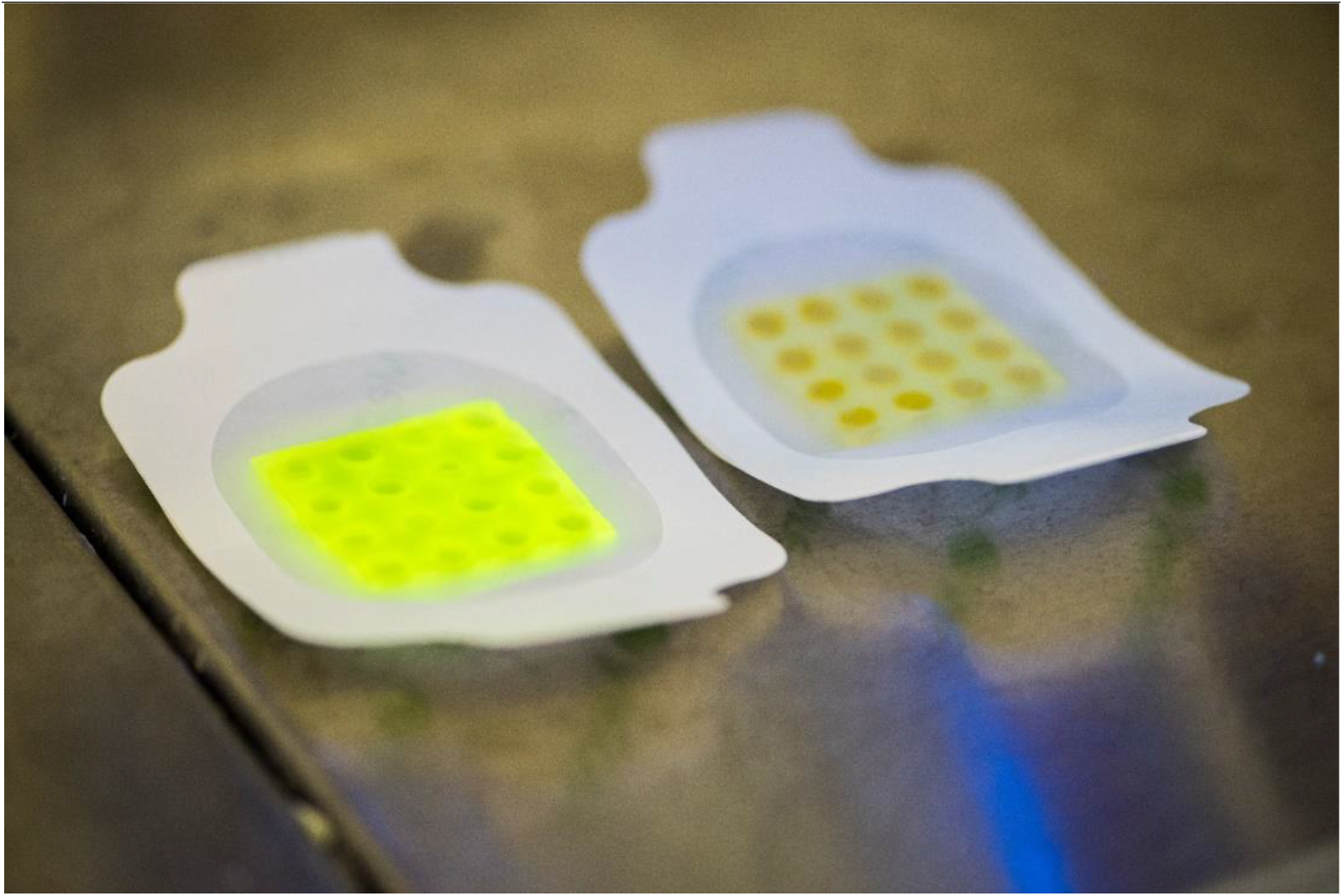
the prototype diagnostic dressing. Left hand side, activated by bacterial virulence factors

In this study, wound dressings removed from consenting patients were couriered to the University of Bath laboratory (within 24 h), where they were placed in phosphate buffer and vortexed to physically remove bacteria. The resultant bacterial suspension was inoculated onto nanoporous polycarbonate membranes (25 mm diameter) placed on an agar plate containing Luria Bertani (LB) broth and a biofilm grown for 24 hours.^7^ After 24 hours the prototype dressing was placed onto the biofilm and after a further 24 hours any colour change of the dressing, caused by the release of carboxyfluorescein from the vesicles was recorded and photographed (figure 2)

**Figure 2:**
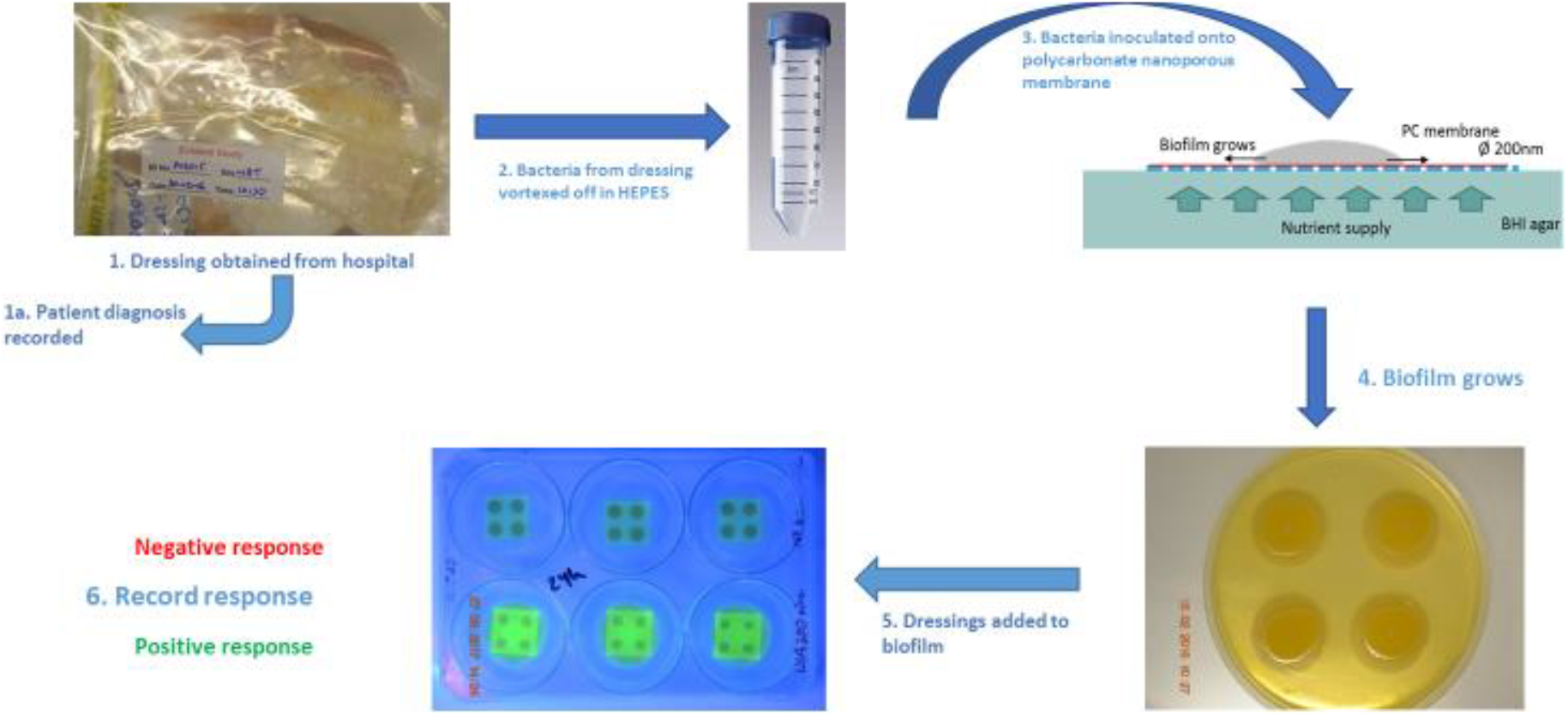
schematic of study approach: Dressings removed from the patient would be anonymised, vacuum packed and shipped to the University of Bath. On arrival they would be immediately vortexed in HEPES buffer to physically remove bacteria and spread on a nanoporous membrane and grown for 24 hours. Dressings would be placed on the polycarbonate membrane and response photographed after 24 hours.

#### Statistical analysis

The baseline demographic and clinical characteristics of participants were recorded.

Cross tabulation of the index test results against those with the three clinical wound signs (reference standard for BWI) were used to estimate the sensitivity of the technology (proportion of participants with BWI) who had a positive test (switch-on), specificity (proportion without BWI who had a negative test), the positive predicted value (proportion of samples positive for the test that were also positive for BWI) and negative predictive values (proportion of negative tests that were negative for BWI) of the test. Confidence intervals around these estimates of accuracy were calculated using the exact binomial method.

##### Sample size

The sample size was set to provide 95% confidence intervals that had two tailed width of no more than 20% (10% in either direction) for a parameter value of 85% for the accuracy statistic with the smallest quantity of data. A sample size of 58 subjects was required in those with BWI to produce a 95% confidence interval of less than 20% where the sensitivity gave a percentage of 85% or more. In order to produce a sample of 58 infection-positive patients where only 20% are positive, and the data set is only complete in 90% of the patients, the required number of patients with diagnosed infection was calculated to be (58*5/.9)=322.

To produce the same level of statistical precision, a sample size of 58 true negatives were needed and a total sample of 100 controls was required. The statistic of sensitivity to detect BWI included all those who had a diagnosed wound infection. The technology specificity was calculated from the negative controls (no wound infection ever diagnosed).

### Calculation of sensitivity, specificity, accuracy and area under ROC curve

Sensitivity and specificity were calculated by calculation of number of true positive (TP), true negative (TN), false positive (FP) and false negative (FN) test outcomes from the dressing.

ROC curves showing the relationship between clinical sensitivity and specificity were calculated. ROC curves are used in clinical research to choose the most appropriate cut-off for a test. The area under the ROC curve (AUROC) was used as a criterion to measure the test’s discriminative ability. AUROC of 0.5 was used to suggest no discrimination, 0.7 to 0.8 was considered acceptable, 0.8 to 0.9 considered excellent, and more than 0.9 was considered outstanding.

#### Dressing switch-on related to BWI

The correlation between dressing switch-on and diagnosed BWI (peri-wound redness; wound pain; local wound temperature) was calculated in terms of dressing/technology sensitivity.

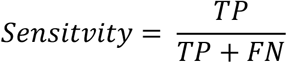

TP = number of true positives; FN = number of false negatives

The dressing specificity was calculated from the “no infection diagnosed” control group (n = 143)

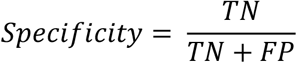

TN = number of true negatives; FP = number of false positives

The area under the Receiver Operator Curve (ROC) was calculated.

Test accuracy was calculated from:

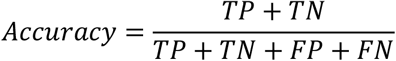

Results are summarised in Table 2.

## Results

### Population demographics

A total of 344 patients were recruited to the study across the three hospital sites. Patients lost to the study are reported in Figure 2. Demographic characteristics of the recruited patients are shown in Table 1. There were 145/288 (50.3%) patients diagnosed with BWI. The mean age of participants with diagnosed infection, and controls, was similar (38 vs. 36 years) and the majority (76%) of participants were adults. A majority (56%) of participants were male with a greater percentage in the diagnosed infection group. For participants where ethnicity was recorded, 84% were white, which was similar across the groups. Just under 10% of ethnicity data were missing. Comorbidities were reported more commonly in the diagnosed infections group than in the controls (37% vs 16%).

**Table 1:** Demographic characteristics of study participants by whether or not infection was diagnosed.

|  | Burn wound infection (n=145) |  |  | Controls (n=143) |  |  |
| --- | --- | --- | --- | --- | --- | --- |
| Characteristic | N | mean or n | s.d. or % | N | mean or n | s.d. or % |
| Age (years) | 145 | 37.9 | 25.7 | 143 | 35.5 | 24.8 |
| Adult (%) | 145 | 113 | 77.9 | 143 | 105 | 73.4 |
| Male (%) | 145 | 83 | 57.2 | 143 | 79 | 55.2 |
| Ethnicity (%) | 125 |  |  | 135 |  |  |
| White |  | 105 | 84.0 |  | 113 | 83.7 |
| Black |  | 2 | 1.6 |  | 7 | 5.2 |
| Asian |  | 10 | 8.0 |  | 8 | 5.9 |
| Mixed/other |  | 8 | 6.4 |  | 7 | 5.2 |
| Comorbidity (Yes vs. No) | 145 | 54 | 37.2 | 143 | 23 | 16.1 |

### STARD Diagram

The STARD diagram is shown below (Figure 3). 311 patients were recruited, with 12 being excluded from analysis due to late sample delivery or sample contamination and 11 further patients were lost to follow-up. The control group (n = 143) of patients with clean burn wounds and no signs of infection were used to test the dressing specificity. Th diagnosed BWI group (n = 145) were evaluated for correlation between dressing switch on and three clinical signs of BWI (Reference standard). In total, dressings from 84 (59%) patients in the control group showed no switch on (true negatives); dressings from 99 (68%) patients in the diagnosed BWI group showed switch-on (true positives). Six dressings from the BWI group and five from the control group gave inconclusive results.

**Figure 3:**
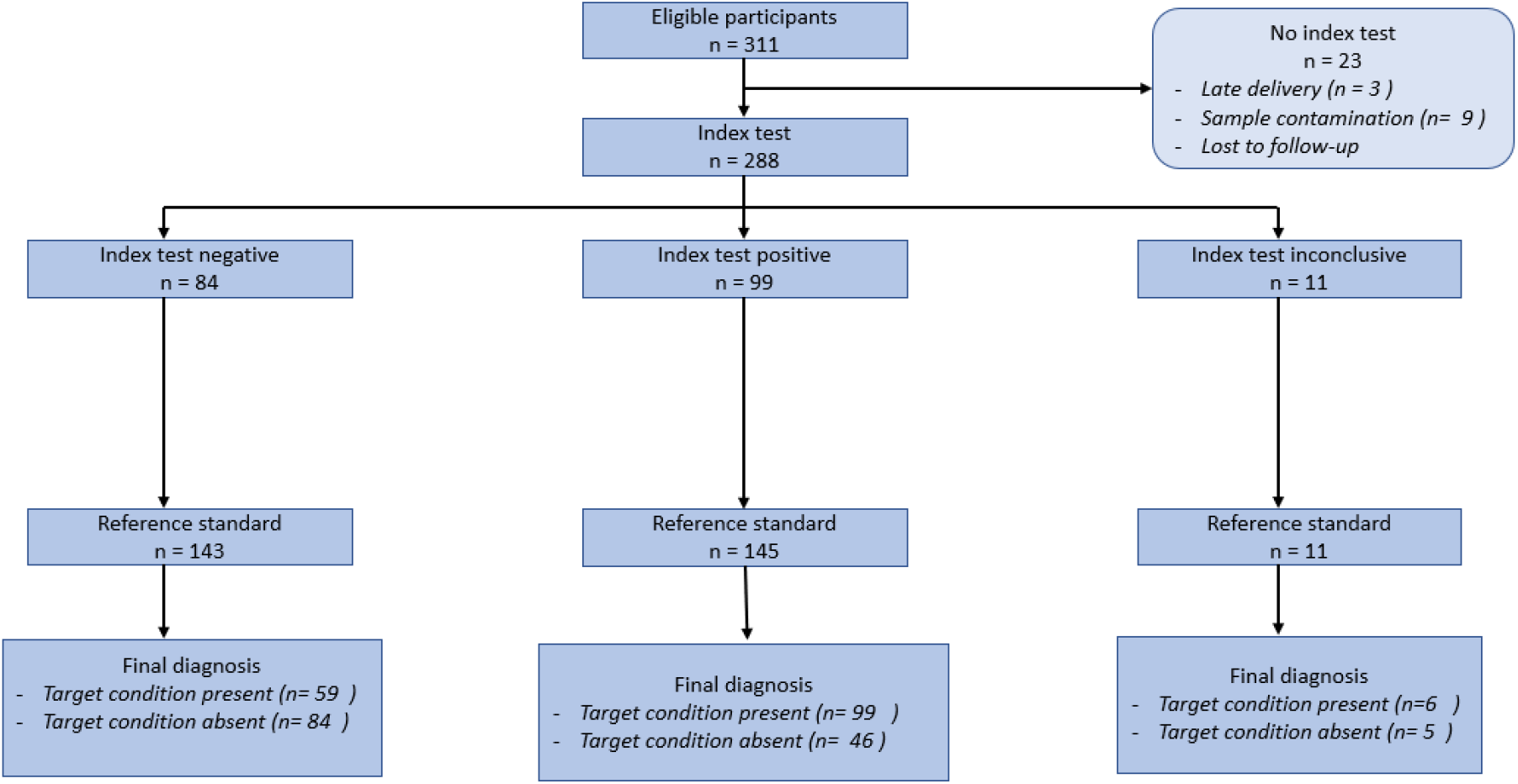
STARD diagram

The dressing ‘switched on’ in 68% of patients with diagnosed BWI using the reference standard (Table 2). The lower confidence limits were above 50% with an upper limit above 70%. Using the controls, the proportion of dressings that did not switch on was 58.7%; lower limit of 95% CI is close to 50% and upper limit is below 70%.

**Table 2:** Agreement between dressing switch-on and symptoms/signs of increased local pain, wound hotness and peri-wound redness

| Performance of dressing | Point estimate | 95% confidence interval | Area under ROC curve (95% confidence) |
| --- | --- | --- | --- |
| Sensitivity (BWI) | 68.0% | (53.3%, 80.5%) | 0.634 (0.5175; 0.737) |
| Specificity (controls only) | 58.7% | (50.2%, 66.9%) |  |

Bacterial species present in the patients wounds are presented in Table 3.

**Table 3:** Bacterial wound presence

| Species detected by routine microbiology | No. of patients in which this occurred |
| --- | --- |
| *Achromobacter xylosoxidans* | 1 |
| *Acinetobacter baumannii* | 3 |
| *Acinetobacter calcoaceticus* | 1 |
| *Acinetobacter iwoffii* | 1 |
| *Acinetobacter nosocomialis* | 1 |
| *Aeromonas* | 1 |
| Alpha haemolytic streptococcus | 5 |
| Anaerobes | 2 |
| *Bacillus Cereus* | 8 |
| *Bacillus species* | 2 |
| *Beta haemolytic streptococcus* | 1 |
| *Beta haemolytic streptococcus A +ve* | 3 |
| *Beta haemolytic streptococcus B* | 6 |
| *Beta haemolytic streptococcus C* | 5 |
| *Beta haemolytic streptococcus G* | 7 |
| *Candida Parapsilosis* | 1 |
| *Candida species* | 4 |
| *Citrobacter koseri* | 1 |
| *Coliform* | 7 |
| *Corynebacterium striatum* | 1 |
| diph colif anaero | 1 |
| Diphtheroid | 8 |
| *Enterobacter Cloacae* | 7 |
| *Enterobacter cloacae complex* | 2 |
| Enterococcus spec. | 9 |
| *Escherichia coli* | 2 |
| *Klebsiella oxytoca* | 3 |
| *Moraxella catarrhalis* | 2 |
| Normal skin flora | 10 |
| oxidase neg Gram Negative Bacilli | 1 |
| *Pantoea agglomerans* | 3 |
| *Pasteurella stomatis* | 1 |
| *Proteus mirabilis* | 3 |
| *Pseudomonas* sp | 1 |
| *Serratia Maracens* | 3 |
| *Serratia species* | 1 |
| *Staphylococcus aureus* - Coagulase negative | 19 |
| *Staphylococcus aureus* - Methicillin Resistant | 1 |
| *Stenotrophomonas maltiphillia* | 2 |
| *Streptococcus* - Non haemolytic | 1 |
| *Streptococcus agalactiae* | 3 |
| *Streptococcus B* | 2 |
| *Streptococcus dysgalactiae* | 8 |
| *Streptococcus pneumoniae* | 3 |
| *Streptococcus pyogenes* | 2 |
| Streptococcus species | 23 |

## Discussion

Clinicians treating patients with acute wounds need a point of care (PoC) diagnostic that clearly indicates and alerts to clinical infection requiring treatment. Current techniques for diagnosis of BWI include use of non-specific symptoms and signs of inflammation, presence of bacteria (which may indicate mere colonisation) and retrospective response to antibiotics (which could mean simple resolution of inflammation). With no definitive ability to diagnose wound infection at the bedside, the clinician is obliged to treat for infection with wound cleaning, superficial debridement and antibiotics to prevent potential sepsis. This results in increased treatment burden and pain for the patient, dressing changes (which impact negatively on healing and promote future scarring), increased unnecessary use of antibiotics (risking an increase in bacterial resistance) and increased healthcare costs.

In order to assess a new technology for PoC detection of BWI, a reference standard is required. As it is not possible to test the prototype technology with an evidence-based reference standard, the three most important wound symptoms/signs (indictors) for infection, chosen by consensus methods in recently published work, were used.^5^ These were: increase in wound pain and temperature and peri-wound redness. Although a clear limitation to a study like this, the use of an expert-agreed reference standard is accepted methodology for diagnostic accuracy studies^9^. Using this methodology, the technology demonstrated a 68% sensitivity and 59% specificity. PoC tests for infection have varying sensitivities and specificities, most of which improve with technical development. As a comparison, some of the early lateral flow tests for the SARS-CoV-2 Antigen evaluated in the autumn of 2020 by the UK governments Scientific Advisory Group for Emergencies, such as the Innova tests system, had sensitivity (true positive rate) of 48.89% (95% CI 33.7% to 64.23%) and a specificity (true negative rate) of 99.93% (95% CI 99.76% to 99.99%), using PCR as a gold standard.^8^ The sensitivity and specificity of the diagnostic tested in the EVIDEnT study mean that the dressing would be less useful for ruling out infection, but more useful for detecting wound infection that requires treatment, and could be used for this purpose without dressing removal - thus improving patient experience and decreasing healthcare costs.

### Study limitations

There are a number of limitations to this study. There are inherent problems in designing a laboratory-based model for testing a prototype dressing technology. The ideal model is the patient wound, but without safety and proof of concept data, testing the dressing directly on the patient is not possible. The rationale of the approach discussed here was to try to re-create the patients wound biome in the laboratory, such that it had a high degree of fidelity with the bacterial population of the original wound. The principal problem with this approach is that the laboratory model will not be an exact facsimile of the original wound biome: in the model, some wound bacteria will not grow or grow very slowly; whilst culturable bacteria may grow at different rates than in the wound - resulting in different proportions of (for example) *Staphylococcus aureus* to *Pseudomonas aeruginosa* in the model than in the original wound biome. The sample size of the study was set for a given precision (confidence interval width). The lower than expected sample size resulted in lower precision. A larger study would increase precision and reliability of the results.

## Conclusion

The results of this *ex vivo* study testing the diagnostic accuracy technology (smart dressing) against wound samples from patients with and without burn wound infection, assessed against a reference standard, showed good sensitivity and moderate specificity.

## Data Availability

Additional data available through Dryad repository: DOI https://doi.org/10.5061/dryad.79cnp5htr

## Acknowledgements

The authors thank The Annette Charitable Trust, Engineering and Physical Sciences Research Council (EPSRC) Grant, references: EP/R51164X/1 and Medical Research Council Grant: MR/N006496/11 and MC_PC_141232.

